# Endpoint PCR Detection of Sars-CoV-2 RNA

**DOI:** 10.1101/2020.07.21.20158337

**Authors:** Samuel Moses, Claire Warren, Phil Robinson, Jon Curtis, Steve Asquith, John Holme, Nisha Jain, Keeley J Brookes, Quentin S. Hanley

## Abstract

Quantitative real-time PCR methods have been used to perform approximately 278 million tests for COVID-19 up to mid-July 2020. Real-time PCR involves a rate limiting step where the samples are measured *in situ* during each PCR amplification cycle. This creates a bottleneck limiting scalability and as a consequence reducing access to inexpensive reliable testing at national and international scales. We investigated endpoint PCR for the qualitative detection of SARS-CoV-2 sequences on synthetic RNA standards and hospital patient samples. The endpoint PCR detection limit is constrained only by the stochastics of low copy numbers and reliably detected single copies of synthetic RNA standards. On a set of 30 patient samples, endpoint PCR found one additional positive sample and was able to confirm an indeterminate sample as negative. These results were found using 4 μl reagent and 1 μl of sample representing an 80% reduction in required RNA extract input and PCR reagent volumes relative to the NHS protocol (20 μl reagent and 5 μl sample). These results indicate that endpoint PCR should be the method of choice for large scale testing programmes. Based on the experience from ultra-high throughput genotyping efforts a single workflow using 384-well plates has similar PCR capacity (250 Million) to that required for all testing done worldwide during the first 7 month of the pandemic.

## Introduction

COVID-19 has emerged rapidly from a few cases in Wuhan, China,^1–3^ to a global pandemic caused by the SARS-CoV-2 virus which is believed to have jumped to humans from an animal host and is one of a host of betacoronaviruses affecting a wide range of animals.^4–7^ This family of pathogens was considered a pandemic threat before the current international crisis began and has defied nearly all attempts to control or eliminate it due to a combination of high infectiousness, undetected carriers and silent transmission.^8–10^ Improved mass scale methods for detecting viral RNA would aid population surveillance for SARS-CoV-2 and put the world in a stronger position for fighting this and other viral diseases.

Real-time PCR is the current standard recommended by the World Health Organisation (WHO) globally for detection of SARS-CoV-2 RNA. This was accepted under emergency regulations in many countries and is now moving to more permanent status. This dominance of testing emerged during the early stages of the pandemic via a WHO listing of diagnostic protocols for COVID-19.^11^ At last update, this included seven protocols developed in China, France, USA, Japan, Germany, Hong Kong, and Thailand targeting a range of SARS-CoV-2 sequences.^11,12^ Each protocol used real-time PCR reflecting the availability of instrumentation and expertise in the labs creating the protocols. The universality of real-time PCR methods in the early tests developed for SARS-CoV-2 created a preference in favour of this readout method which has persisted.

Although real-time quantitative PCR is a standard method in diagnostic as well as research labs and universities,^13^ it can be argued that real-time fluorescence signal reading and monitoring is not always required for tests that require only a qualitative binary (Yes/No) interpretation for both surveillance as well as diagnostic screening purposes. Industrial scale qualitative PCR using rapid water-bath temperature cycling followed by endpoint readout was described first in 1993 with a scale of 46,000 parallel reactions in 384-well plates.^14,15^ This progressed to 1536-well formats by 2005^16^ and capacity has since grown such that a single lab using endpoint PCR can carry out 1.5 million PCRs per day at reasonable cost for absence/presence SNP genotyping.^17–19^ A single 384-well plate industrial PCR system is capable of 800,000 samples per day (146 M per 6 months) and with full 24/7 could exceed this.

Endpoint PCR and low-cost reagent mixes underpin a cost-saving revolution in genetics such that genotyping is now less costly than phenotyping.^20–22^ As an example, the KASP endpoint assay system underpins hundreds of papers since 2019 (c.f. ^19,22,23^). There have been thousands of papers in the last decade proving the robustness of end-point PCR on over 400 genomes particularly for important food crops. Cost and performance comparisons have appeared in the literature and industrial scale PCR cost has been well below US$1/data point for several years and costs as low as US$0.064/data point documented.^22–24^ A similar cost and scale revolution in SARS-CoV-2 testing would enable high-quality monitoring in even the poorest of countries.

Scale limitations create a range of sampling, storage and processing issues. For example, false negatives (FN) have been an ongoing problem with FN rates as high as 50% reported.^25,26^ With limited capacity, retesting of patients or samples is not always feasible. The ultra-high throughput capabilities of endpoint PCR technologies provide an opportunity to correct this. Further, at least in the UK, the cycle threshold and copy number values provided by real-time PCR are not being used to inform clinical practice in any way and, to our knowledge, this is true worldwide. Therefore, the extra unused information provided by real-time methods is creating an unnecessary and costly bottleneck. Real-time PCR is providing the same Yes/No result for the presence/absence of a short length nucleotide sequence that endpoint PCR can provide at lower cost and higher throughput. Endpoint PCR has been demonstrated in forensic science^27^ and for detection of other infectious diseases such as Hepatitis B,^28^ Ebola,^29^ and HIV^30^ with some authors indicating endpoint methods are more sensitive than real-time.^30^

Here, we investigated endpoint PCR as an alternative to real-time PCR for COVID-19 (SARS-CoV-2 RNA) testing. We compare the two techniques on authentic positive samples collected at Kent hospitals during the pandemic and report detection limits using hospital patient samples and a dilution series using synthetic SARS-COV-2 sequences.

## Materials and Methods

### Sensitivity Standards

Synthetic RNA controls containing nominally 10^6^ copies/μl of the Twist Bioscience, Control 2 sequence (GISAID Wuhan-Hu-1; Genbank ID MN908947.3; Twist Bioscience) were diluted to a starting concentration of 100,000 copies per μl using the mass information provided by the manufacturer. Standards were prepared by serial dilution to obtain 10,000, 1,000, 100, 10, 1, 0.1, and 0.01 copies per μl in 0.1mM Te (10mM Tris, pH 8.3, 0.1 EDTA). These were tested using 2 μl aliquots using 10 replicates.

### Samples and extraction

A set of 30 anonymised combined nasopharyngeal & oropharyngeal samples collected from patients presenting to Kent Hospitals Trust with COVID-19 symptoms were considered. Using NHS in-house testing 19 of these samples were positive, 10 negative, and 1 indeterminate. All samples were extracted using either the Promega extraction kit or the Abbott M2000 method. The ensuing Promega extracts underwent PCR by either GeneFinder COVID-19 PLUS Real*Amp* Kit or the Viasure SARS-CoV-2 Real Time PCR Detection Kit. The Abbott M2000 extracts were part of the single-flow closed extraction and PCR analyser process. The specific extraction methods are described below:

#### Promega Extraction

300 μl of nasopharyngeal and oropharyngeal swab sample in Sigma Virocult® MW951S – MWE (Medical Wire) collection medium was inactivated post-collection by adding 300 μl of Promega Lysis Buffer (1:1). 30 μl proteinase K was added to 300 μl of the resulting solution which was then extracted with Viral Total Nucleic Acid Purification Kit (Promega) giving 50 μl of sample for PCR tests.

#### M2000 Extraction

500 μl of nasopharyngeal and oropharyngeal swab sample in Sigma Virocult® MW951S – MWE (Medical Wire) collection medium was combined with 500 μl of Promega Lysis Buffer and 500 μl of the resulting solution was extracted with the M2000 extraction apparatus as specified by the manufacturer resulting in 90 μl of RNA eluate for PCR tests.

The extracted RNA eluates were stored at -80°C prior to analysis.

### PCR

The anonymised samples’ RNA eluates were tested using two protocols. First, the samples were analysed using the Kent Hospitals diagnostic protocols. The Hospital RT-PCR diagnostic protocols used GeneFinder RealAmp kit for RdRp (RNA dependent RNA polymerase), E (envelope), N (nucleocapsid) sequences; Abbott RealTi*m*e SARS-C0V-2 Amplification Reagent kit for the RdRp and N sequences; and Viasure SARS-CoV-2 Real Time PCR kit for orf1ab (open reading frame 1ab) and N sequences. The samples had no specified pre-test selection in terms of which assay was used as all of the assays were validated and verified to be of comparable performance criteria (sensitivity, specificity) set against clinical as well as analytical external controls as part of a multi-option diagnostic pathways for dealing with the SARS-CoV-2 pandemic.

#### Hospital real-time PCRs

Both the GeneFinder RealAmp PCR and the Viasure SARS-CoV-2 Real Time PCR assays used 5 μl of RNA eluate added to 20μl of respective assay’s detection mastermix which then underwent PCR reaction in a 25 μl reaction (96-well plate).

The Abbott RealTi*m*e SARS-C0V-2 assay utilised 15 μl of RNA eluate from Abbott M2000 extraction added to 90μl of assay detection mastermix prior to real-time PCR in the Abbott M2000 analyser.

#### Endpoint PCR

The same RNA eluates from the 30 anonymised samples underwent endpoint PCR as follows. 1 μl of the RNA eluates were run in a final reaction volume of 5 μl of commercial RT-PCR mastermix optimized to facilitate endpoint readout and containing 500 nM ™ROX according to the manufacturer’s instructions (ProbeSure™ Covid19 One Step RT-PCR endpoint mix; COV-1010-1; 3C^**r**^ Bioscience). This mix contains primers and probes for the N1, N2 SARS-CoV-2 genes and and RNaseP control using the sequences: (2019-nCoV_N1-Forward Primer : 5’-GAC CCC AAA ATC AGC GAA AT-3’, 2019-nCoV_N1 Reverse Primer : 5’-TCT GGT TAC TGC CAG TTG AAT CTG-3’, Probe N1 : FAM-ACC CCG CAT /ZEN/ TAC GTT TGG TGG ACC-3IABkFQ; 2019-nCoV_N2 Forward Primer : 5’-TTA CAA ACA TTG GCC GCA AA-3’, 2019-nCoV_N2 Reverse Primer : 5’-GCG CGA CAT TCC GAA GAA-3’, Probe N2 : FAM-ACA ATT TGC /ZEN/ CCC CAG CGC TTC AG-3IABkFQ; RNAse P Forward Primer : 5’-AGA TTT GGA CCT GCG AGC G-3’, RNAse P Reverse Primer : 5’-GAG CGG CTG TCT CCA CAA GT-3’, Probe RNase P : FAM-TTC TGA CCT GAA GGC TCT GCG CG-3IABkFQ. The probes in this mix for N1, N2, and RNase P were obtained from IDT (Product codes: N1 probe: 10006823; N2 probe: 10006826; RNase P probe: 10006829; Integrated DNA Technologies, Inc.) The standards were measured using a duplex mix to test both N1 and N2 in the same well. The sequences were as above except N1 was labelled with ™FAM and N2 with ™HEX. 2 μl of RNA standard was analysed in a final reaction volume of 5 μl (384-well plate).

### Readout and presentation

The endpoint samples and standards were thermally cycled for 50 PCR cycles and quantified using fluorescence readout for ™FAM, ™HEX and ™ROX dyes as needed on a 7900HT (Applied Biosystems) instrument to derive real-time PCR data. The same sample plate was then read with a standard plate reader (Tecan Spark) to generate the endpoint data. The ™ROX signal was used to normalise the acquired data to correct for variations in pipetting across wells.^31^

## Results and Discussion

### Sensitivity tests and standards

To test the sensitivity of the endpoint system replicate (*N* = 10) standards for each gene were measured (Figure 1) giving 20 measurements per concentration. Of the 160 measurements, one at 200 copies/well resulted in an indeterminate result due to a pipetting failure (no ™ROX was observed). No negative controls amplified. The N1 and N2 replicates gave ™ROX normalised values of 0.0501±0.0017 (*N*=10) and 0.1052±0.0032 (*N* = 10), respectively. We considered any value further than 10 sd from the negative controls (0.07 and 0.14) to be detected. Between 20-20,000 copies/well, the genes were detected in all cases except the indeterminate well and formed a tight cluster distant from the negative controls (Figure 1a). Below 20 copies/well, stochastic effects were seen such that the signal levels varied and, as expected, copies were not detected in all wells (Figure 1b, 1c, and 1d). The 2 copies/well standard amplified sufficiently to be detected in 13 out of 20 cases, 0.2 copies/well amplified in 2 out 20 as did 0.02 copies/well. A simulation consisting of 10,000 repeats of 20 draws from a Poisson distribution having means of 20, 2, 0.2, and 0.02 gave means of 20, 17.2, 3.62, and 0.398 wells, respectively, for the number of wells expected to have at least one copy of the sequences. This was in reasonable agreement the 20, 13, 2, and 2 wells with detected genes. The lowest concentration is high relative to the expected mean; however, our simulation gave 2 or more amplifications in approximately 6% of the 10,000 trials. The variability expected from a Poisson process under these conditions can be seen in the histograms produced by the simulations (Figure 1e). These results indicate that the endpoint PCR strategy can detect SARS-CoV-2 genes with excellent sensitivity down to levels where variability is dominated by the stochastics of single copies. The quality of the data makes detection of the genes straightforward and well suited to either manual or automated analysis.

**Figure 1:**
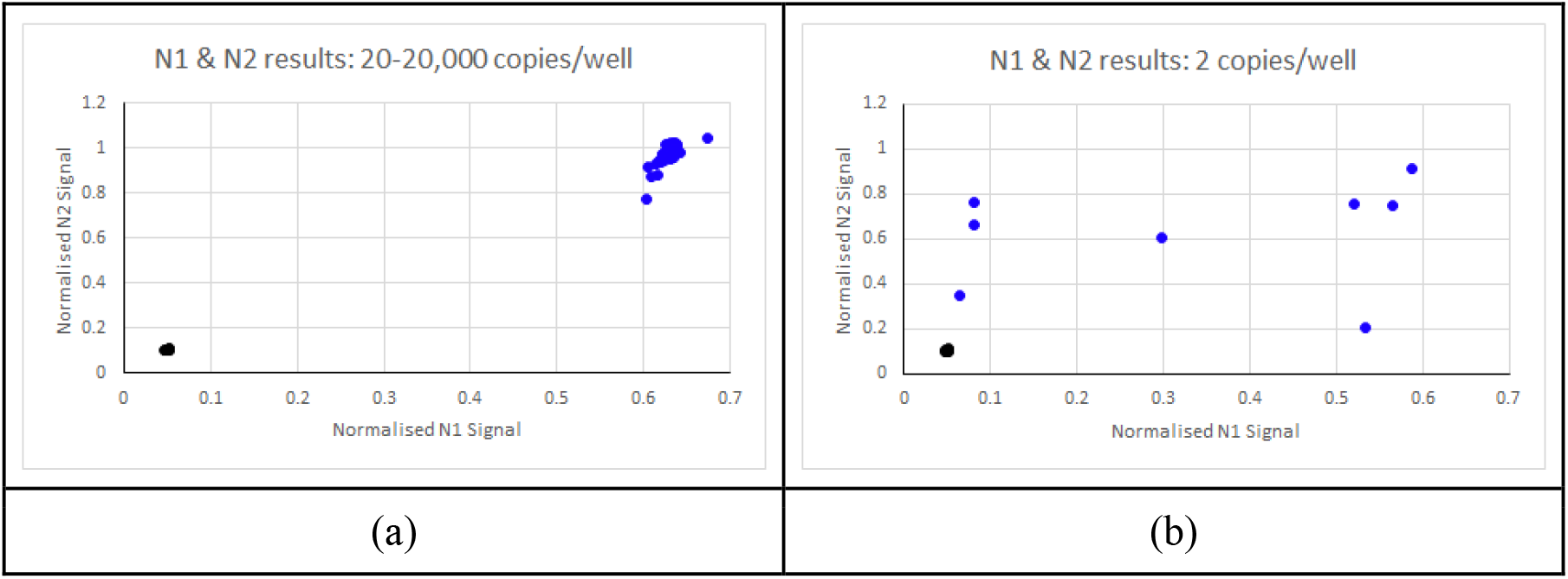
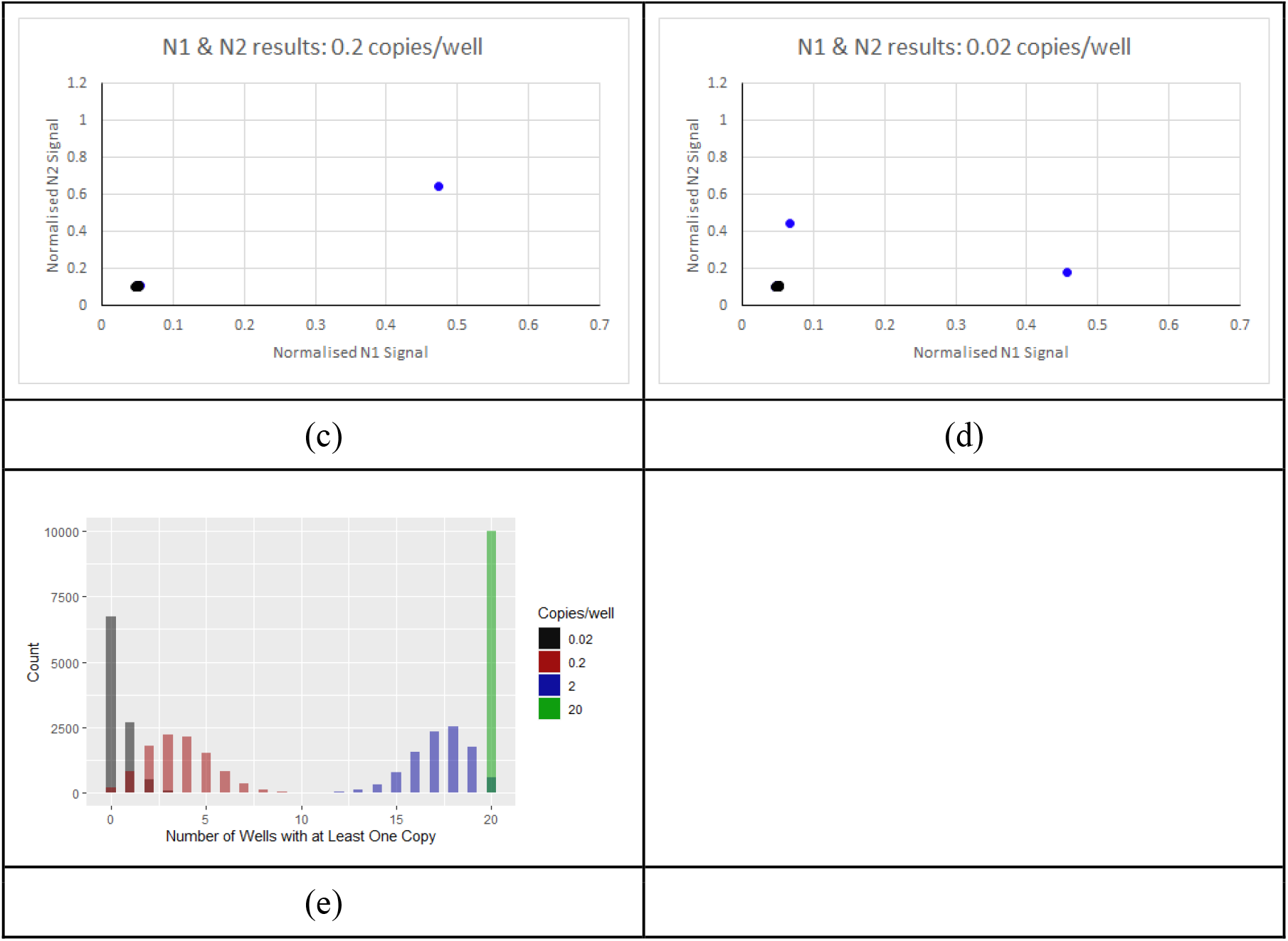
Standard sample measurements and Poisson simulation of 0-20,000 copies/well. Plots show paired ™ROX normalised signals for the two genes in the standard as (N1, N2). The standard with 20-20,000 copies per well (a) gave good discrimination between the standard and controls. Below 20 copies/well stochastic effects were seen (panels b, c, and d). No amplification was observed in the negative controls (black). Panel e shows the results of simulations of a Poisson process corresponding to the expected behavior of 0.02, 0.2, 2, and 20 copies per well.

### Hospital Sample Controls and threshold

To test whether the synthetic material gave an unrealistic view of the sensitivity of the method, we evaluated the RNaseP positive and negative controls using the hospital material. The ™ROX normalised RNaseP measurements gave 1.185±0.036 (*N*=30) and the negative controls 0.1850±0.0070 (*N*=6). Any threshold value above 0.255 (10 standard deviations above the negative control) gives very high confidence that a positive sample has been detected in the hospital samples (Figure 2). This yields a simple yes/no answer. Although the positive samples were slightly lower than the positive controls, they form a tight readily interpretable cluster far away from the negative samples and negative controls. This is a feature of reagent mixes suited to endpoint PCR because they are designed such that the fluorescence intensity reaches the same level independent of the input RNA concentration. This results in simple interpretation that can be easily automated for large scale screening or done manually.

**Figure 2:**
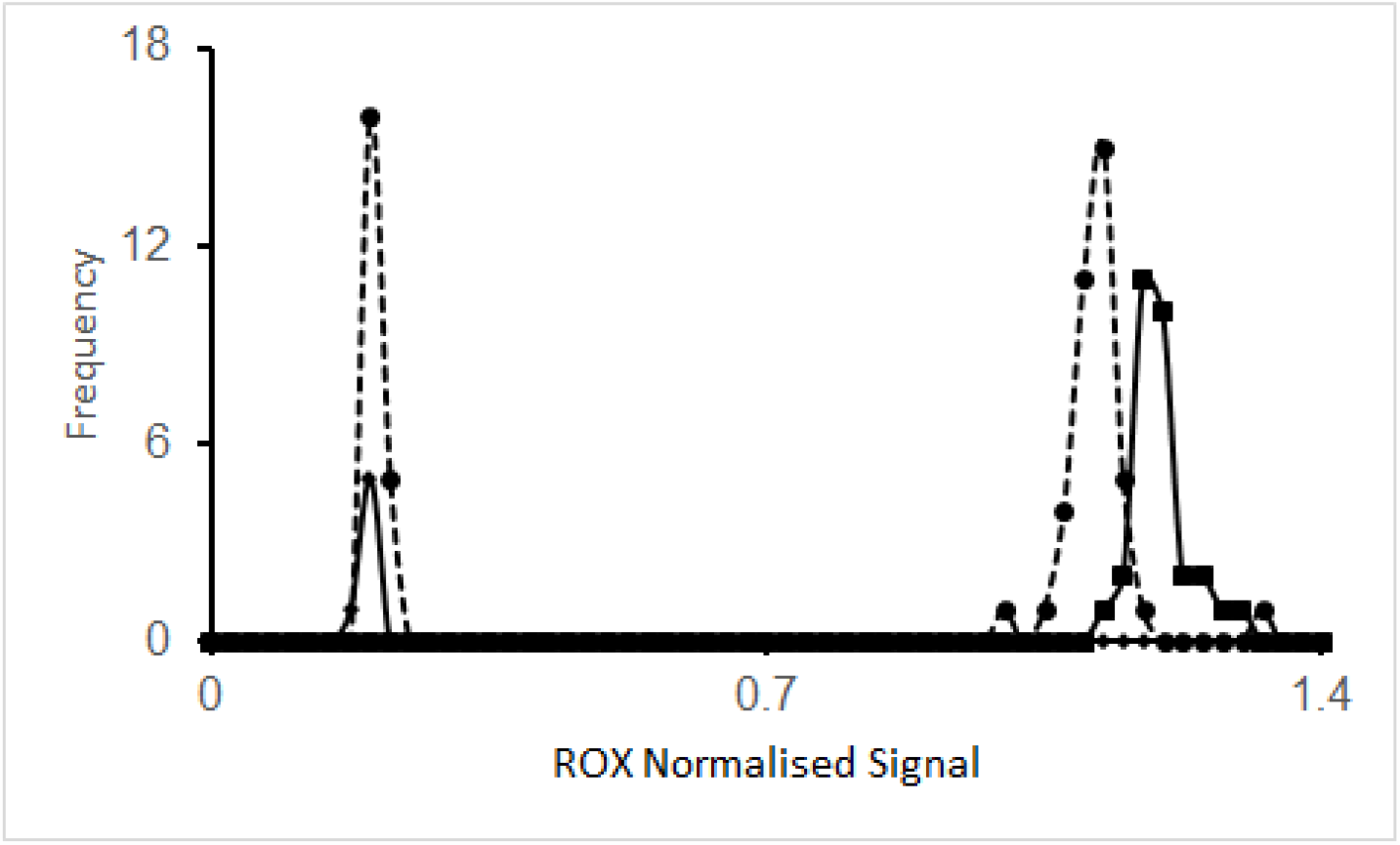
Histogram of ™ROX normalized signals from samples (circles, dashed lines) and positive (squares, solid line) and negative (diamonds, solid lines) controls.

To demonstrate the endpoint method, a set of samples were evaluated that had been previously measured in Kent Hospitals on patients presenting to hospital with COVID-19 symptoms. These samples were measured with RNase P as a positive control (Figure 2) and read using both real-time PCR (Figure 3a and 3b) and endpoint PCR (Figure 3c) for the SARS-CoV-2 N1 and N2 sequences in a reaction volume of 5 μl. The real-time PCR C_T_ values ranged from 12-37 and were comparable to those found by the NHS testing lab (C_T_ from 14-37) in a 25 μl reaction (96 well plate). The decisions when compared to the previous results obtained in the Kent Hospitals Laboratory were nearly 100% concordant. One sample found to be negative by NHS testing was positive for the N1 gene but not N2. This sample is prominent in the endpoint presentation (Figure 3c) as the point near the *x*-axis. This was assigned as a positive. In addition, the indeterminate NHS sample was found to be negative.

**Figure 3:**
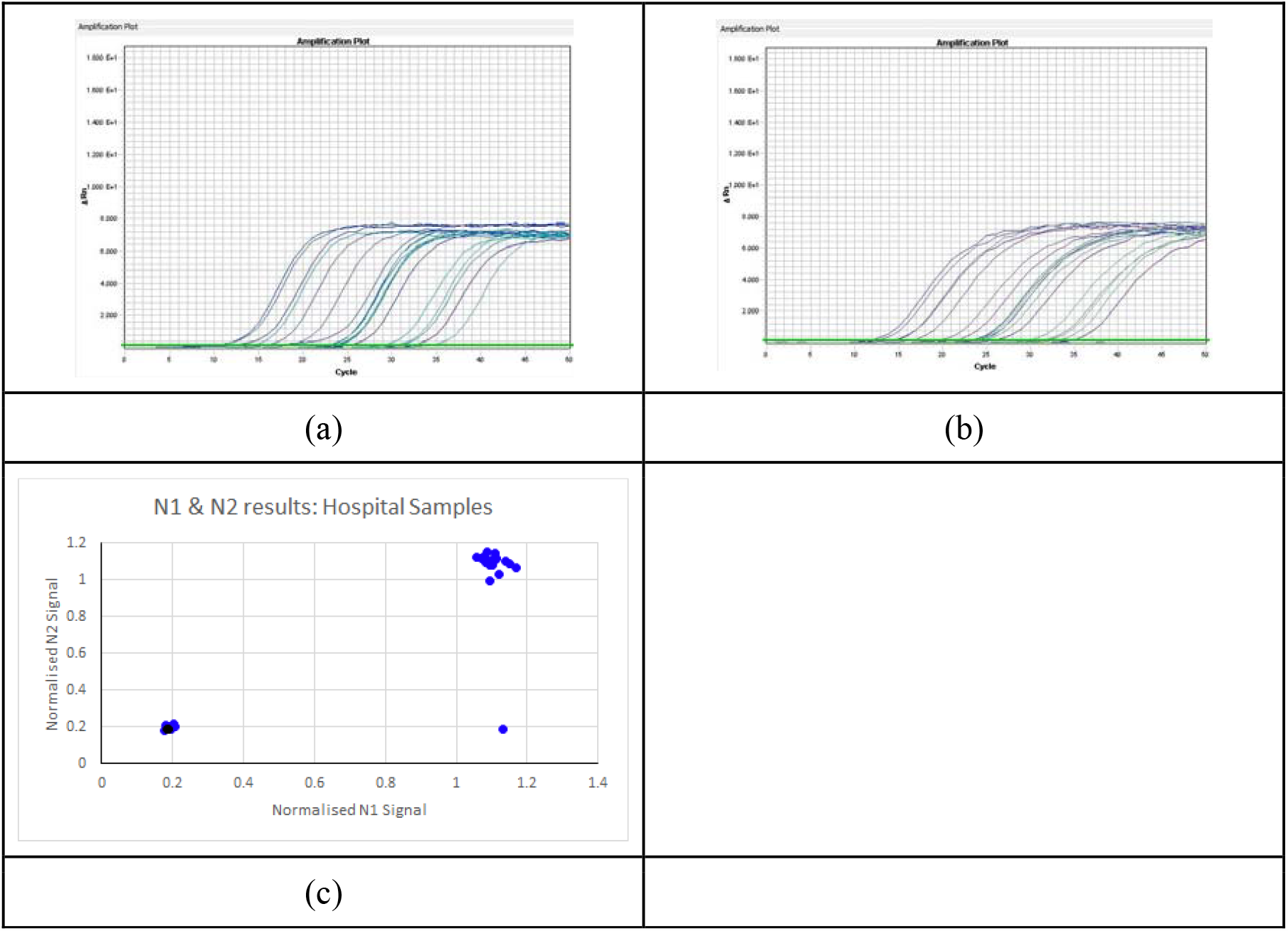
Comparison of real-time and endpoint PCR results for patient samples. Standard real-time PCR traces for the N1 (a) and N2 (b) genes. The endpoint presentation (c) plots the results of two genes normalised to the ™ROX signal as *xy*-coordinates. Black markers represent samples testing negative.

## Discussion and Conclusion

We demonstrated that endpoint PCR is an excellent method for detection of SARS-CoV-2 RNA from patient samples and synthetic standards. Its sensitivity and variability are limited only by the stochastics of low copy numbers. It was able to detect an additional positive sample and resolve an “indeterminate” result as negative. It provides a simplicity of readout giving a clear yes/no answer while removing an equipment bottleneck in testing workflows. This confirms previous work using endpoint PCR for other diseases. Endpoint PCR is inherently more scalable because the thermocycling step is done in parallel prior to readout rather than *in situ*. Real-time PCR limits instrument throughput by monitoring the increased fluorescence as thermocycling takes place. As such, endpoint PCR should be the method of choice for mass testing populations in hospitals, schools, factories, cities, and countries and is readily adaptable to any disease where a sequence is known.

The real-time PCR methods currently in near universal use reflect the equipment and methodology available in the labs that developed the tests for SARS-CoV-2. Real-time methods have struggled to keep up with testing requirements and the widespread use of 96-well plate technology is not the best use of valuable reagents under these conditions. Up to the time of writing (15 July 2020), a total of 278,143,685 tests have been reported worldwide. A single endpoint PCR workflow using 384-well plate formats operating 140 plates an hour 24/7 could have provided 220M (million) RNA PCR results over 198 days. This capacity estimate is meant as an indicator of the scale that has been realised industrially on DNA on difficult samples (plants). Two to three such platforms will give robustness and provide opportunity for replicate analysis. Until vaccines are available or clinical interventions improve, finding and isolating people with COVID-19 is the only way to fight the disease. Endpoint PCR can realistically provide the PCR capacity needed at country scale and reasonable cost.

Existing real-time PCR labs can switch to endpoint PCR by switching chemistries while using existing equipment which can be replaced as needed. Many countries and industrial organisations may already have much of the required equipment lowering the overall costs and the up-front cost of an installation with 800,000 sample/day capacity is circa £2.3 million. This is insignificant in comparison to the growing loss of life and the economic damage done by the SARS-CoV-2 virus. Even the richest countries will be unable to withstand further extended periods of reduced economic activity leaving an impossible task balancing health and economic hardship. Endpoint PCR methods can provide country scale PCR capacity for surveillance including multiple retesting and the infrastructure can be adapted to a wide range of diseases as needed.

## Data Availability

The data presented and a concordance table are included as supplementary materials.

## Data Availability

The data presented and a concordance tables are available as supplementary materials.

## Contribution

This study was designed by J. Curtis, P. Robinson, S. Moses, K. Brookes and Q. Hanley. All authors contributed to the preparation of the manuscript. The lab experiments and analysis measurements presented were performed by C. Warren, N. Jain, S. Asquith and J. Holme. Final editorial decisions were made by J. Curtis, K. Brookes, S. Moses and Q. Hanley.

